# Predicting the end of Covid-19 infection for various countries using a stochastic agent-based model taking into account vaccination rates and the new mutant

**DOI:** 10.1101/2021.04.22.21255941

**Authors:** Manfred Eissler

## Abstract

Using a stochastic, agent-based model, the course of infection since the first occurrence of a Covid-19 infection is simulated for various countries, taking into account the new, more infectious mutant and the vaccinations. The simulation shows that the course of infection for the United Kingdom (UK) and Israel is surprisingly good. For the other countries, an end date for the infection can be predicted based on the course of the simulation. For Germany, the course is calculated in a second scenario, assuming a higher vaccination rate.

## Background

In April 2021, the infection figures in Israel and the UK have fallen to very low levels despite the new, more infectious mutant B 1.1.7. The decisive reason is probably the high vaccination rate in these countries, reinforced by lock-down measures. In other countries, the infection figures are rising.

Using real data, an attempt is made to replicate the course of infection in selected countries with a stochastic, agent-based SIR model and to make a prediction for the further course of infection.

Stochastic agent-based models can be used to simulate infection trajectories very well, with the results of such simulations being random variables that vary slightly with each run. Mathematically, this is a Markov process.

## Method

This type of model will be used to replicate the course of infection in Israel and the UK, and simulations will be carried out for other countries, with the aim of predicting the end of the epidemic, taking into account the respective vaccination rates and the increase in the baseline R0 reproduction rate due to the emergence of a new, more infectious mutant.

The model used is described in [1]. It begins with an infected person in the middle and uses two essential elements: first, the stochastic infection of the immediate neighbours in a 2-D plane consisting of 90,000 persons. In addition, the basic reproduction rate R0 can be set. Furthermore, there is a mobility - also stochastically controlled - of the infected individuals. This leads to the emergence of new infection foci. Figure 1 shows an example visualisation after 330 time steps.

**Fig. 1.**
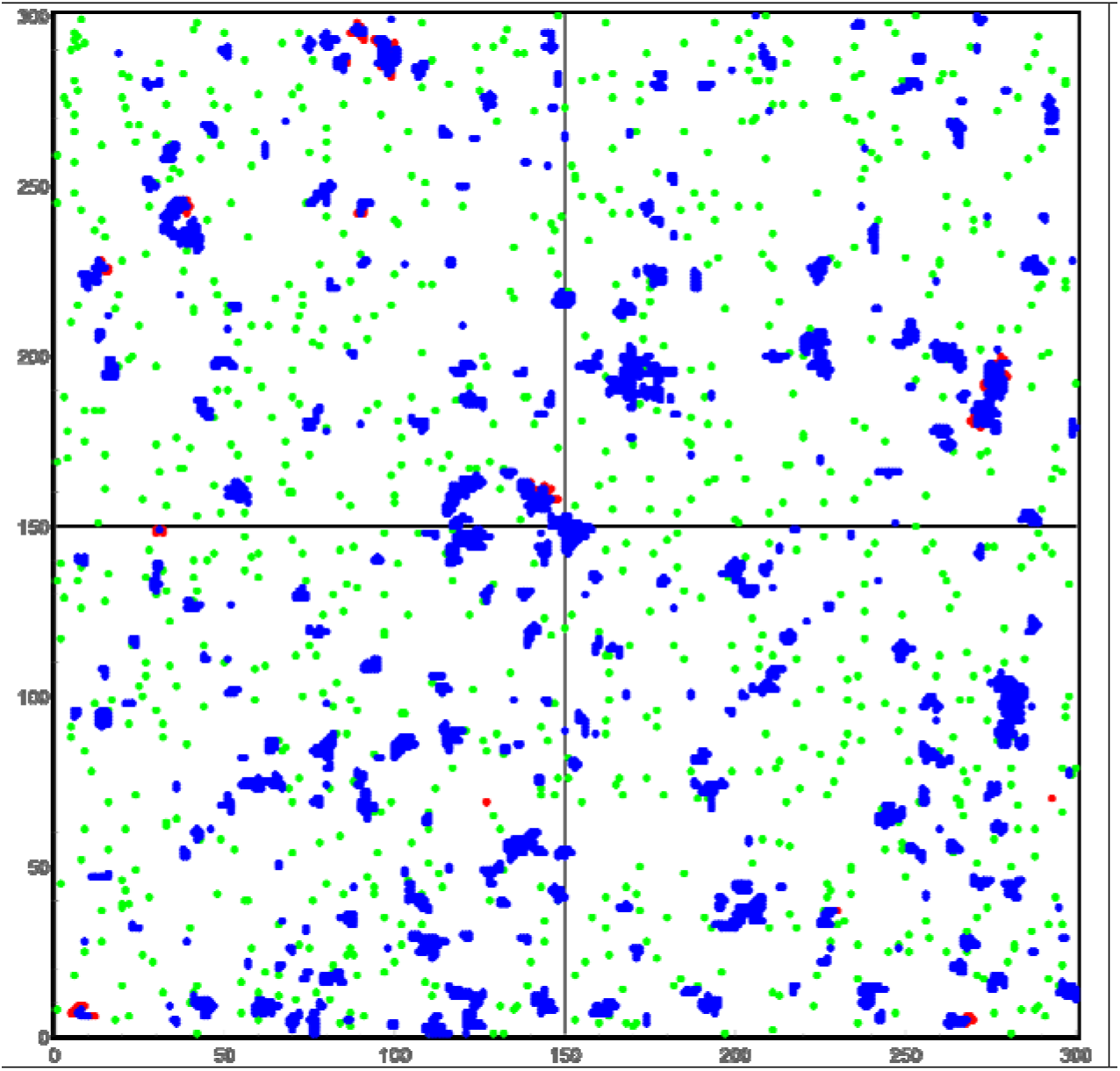
Simulation after 330 time steps. Red Points: Infected, Blue Points: Restored, Green Points: Vaccinated.

For each country, the simulation starts on the date when infections are first listed in the John Hopkins University (JHU) data [2]. The initial R0 value of 2 is reduced by a reduction factor after 10 time steps (Fig.2). This reduction factor is calibrated for each country so that, at the mutant’s start date, the cumulative infected persons in the simulation are higher than the cumulative number of infected persons of the JHU values by a factor that can be interpreted as the estimate of the number of unreported/unidentified cases. These values lie between 1.2 and 2.6 for the countries considered. There is a wide range of assumptions for the actual number of unreported/unidentified cases [3], [4], [5]. With these simple prerequisites, the real course of infection, which often shows a pronounced wave form, cannot be represented up to the point of the “start of the mutant”. However, the values at this point are decisive, since they essentially determine the further course according to a Markov process. Column 4 of Table 1 shows the value of the simulation and the real value of the proportion of infected cumulated for the respective country.

**Tab.1.**

| Country |  | Start Infection | Start Mutant | Start Vaccination | Last Retrieval | End Infection |
| --- | --- | --- | --- | --- | --- | --- |
| UK | Date | 30-Jan-20 | 1-Oct-20 | 8-Dec-20 | 11-Apr-21 | 31-Mar-21 |
|  | Model: Infected cum |  | 0.021 |  | 0.073 | 0.073 |
|  | Reality (JHU): Infected cum |  | 0.008 |  | 0.065 | 0.072 |
| Israel | Date | 20-Feb-20 | 1-Dec-20 | 19-Dec-20 | 11-Apr-21 | 28-Mar-21 |
|  | Model: Infected cum |  | 0.078 |  | 0.109 | 0.109 |
|  | Reality (JHU): Infected cum |  | 0.040 |  | 0.098 | 0.098 |
| Germany 1 | Date | 26-Jan-20 | 7-Dec-20 | 27-Dec-20 | 11-Apr-21 | 6-Jul-21 |
|  | Model: Infected cum |  | 0.036 |  | 0.063 | 0.072 |
|  | Reality (JHU): Infected cum |  | 0.015 |  | 0.036 | ? |
| Germany 2 | Date | 26-Jan-20 | 7-Dec-20 | 27-Dec-20 | 11-Apr-21 | 13-May-21 |
|  | Model: Infected cum |  | 0.040 |  | 0.069 | 0.072 |
|  | Reality (JHU): Infected cum |  | 0.015 |  | 0.036 | ? |
| France | Date | 23-Jan-20 | 7-Dec-20 | 27-Dec-20 | 11-Apr-21 | 17-Jul-21 |
|  | Model: Infected cum |  | 0.047 |  | 0.084 | 0.102 |
|  | Reality (JHU): Infected cum |  | 0.036 |  | 0.079 | ? |
| Italy | Date | 30-Jan-20 | 7-Dec-20 | 27-Dec-20 | 11-Apr-21 | 5-Jul-21 |
|  | Model: Infected cum |  | 0.039 |  | 0.071 | 0.084 |
|  | Reality (JHU): Infected cum |  | 0.029 |  | 0.062 | ? |
| Netherlands | Date | 26-Feb-20 | 23-Nov-20 | 6-Jan-21 | 20-Apr-21 | 14-Jul-21 |
|  | Model: Infected cum |  | 0.034 |  | 0.084 | 0.090 |
|  | Reality (JHU): Infected cum |  | 0.029 |  | 0.084 | ? |

From the country-specific date of the first appearance of the new, more infectious mutant B.1.1.7, R0 increases. This increase depends not only on the not precisely known infectivity of the new mutant, but also on country-specific infection control measures. This increase is modelled in such a way that the country-specific course of infection can be represented as best as possible. With these calibrations, the model can then simulate the further course of infection until the end of the infection.

## Results

Figure 2 shows the result of the simulation for the United Kingdom (UK). The upper blue wave shows the course without vaccination. Within 8 months after the first appearance of the new mutant, 89 percent of the population would be infected due to the rising R0 value caused by the increased infectivity of the new mutant (and possible relaxation of restrictions). This is certainly an unrealistic scenario, as any government would take massive countermeasures in the event of such a progression. The red line shows the simulated course if vaccinations are carried out starting a certain point in time. The vaccination rates used were taken from the data in [6]. In Figure 2 and the subsequent diagrams for the other countries, this vaccination rate is shown schematically over time (with a reduction factor - green line).

**Fig.2.**
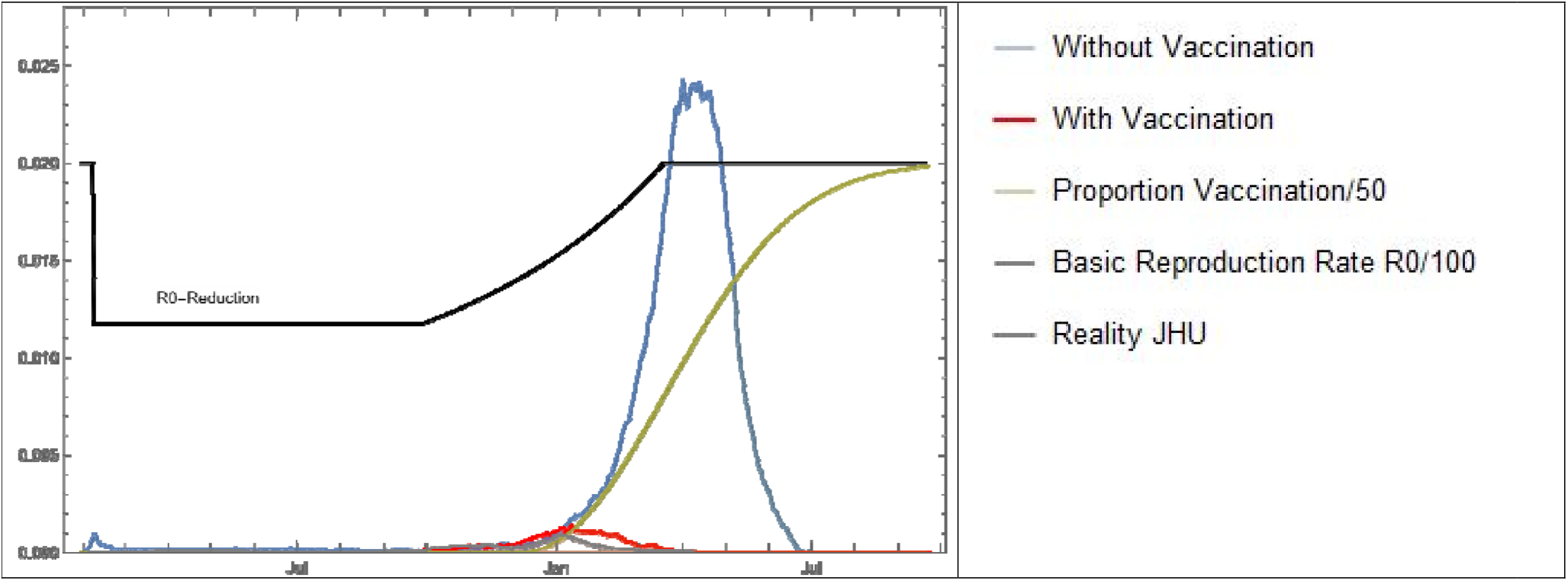
Course of infection for the United Kingdom (UK) without and with vaccination. Schematic representation of R0 and the vaccination rate.

Figure 3 a-g shows enlarged curves of the number of infected persons for different countries: red represents the results of the simulation, while grey is the real curve with the values of the JHU. The calibrated course of R0 and the temporal course of the vaccination rate are also shown. Except in Fig.3d, the respective vaccination rates correspond to the real values up to the last call-up day (Table 1, column 6), which were taken from [6,7]. The data beyond this date were extrapolated.

**Fig.3a.**
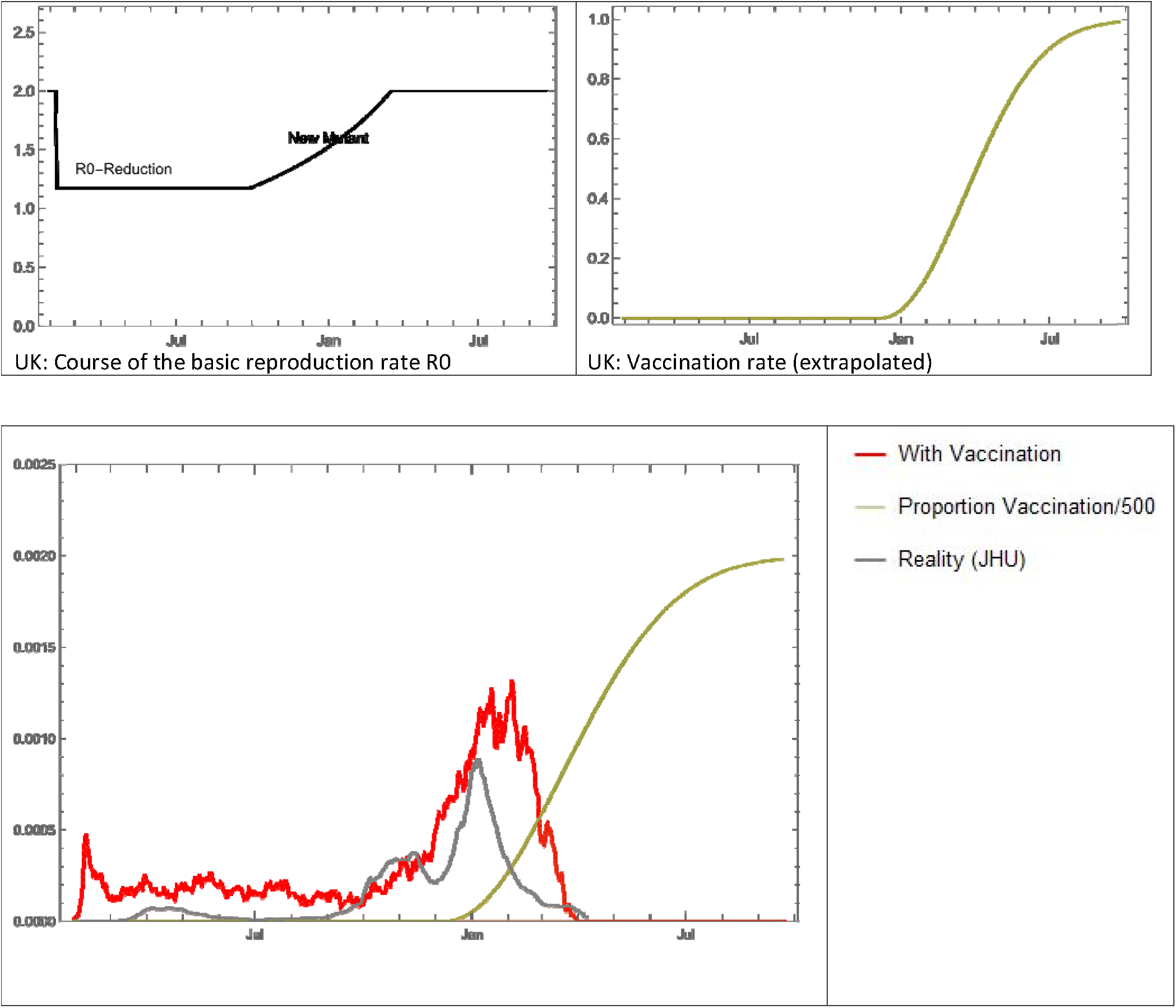
UK: The red curve shows the simulation of the infection with vaccination, the grey curve shows the real course of the infection (Moving Average 7 Days), green: schematic the vaccination rate.

**Fig.3b.**
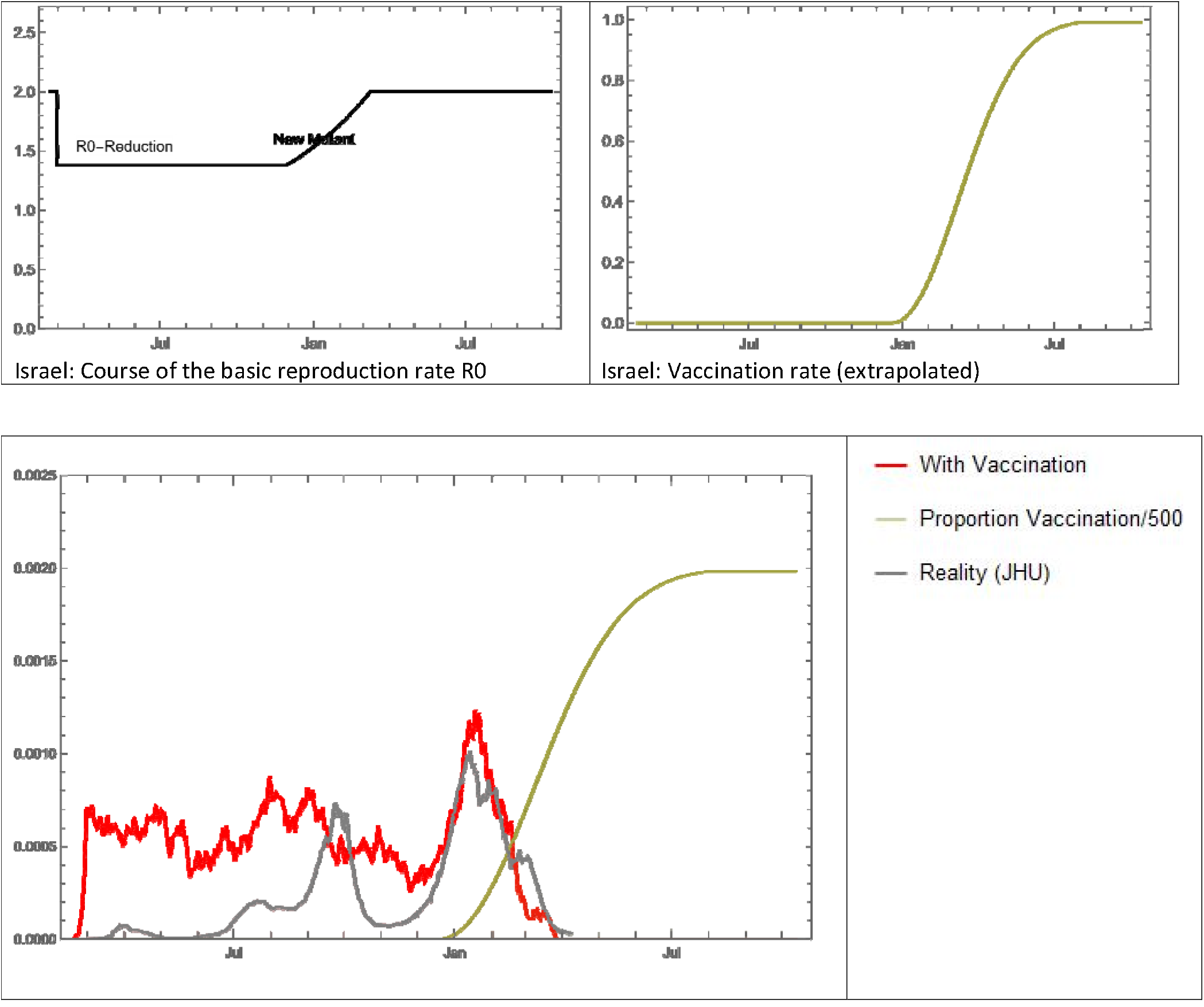
Israel: The red curve shows the simulation of the infection with vaccination, the grey curve shows the real course of the infection (Moving Average 7 Days), green: schematic the vaccination rate.

**Fig.3c.**
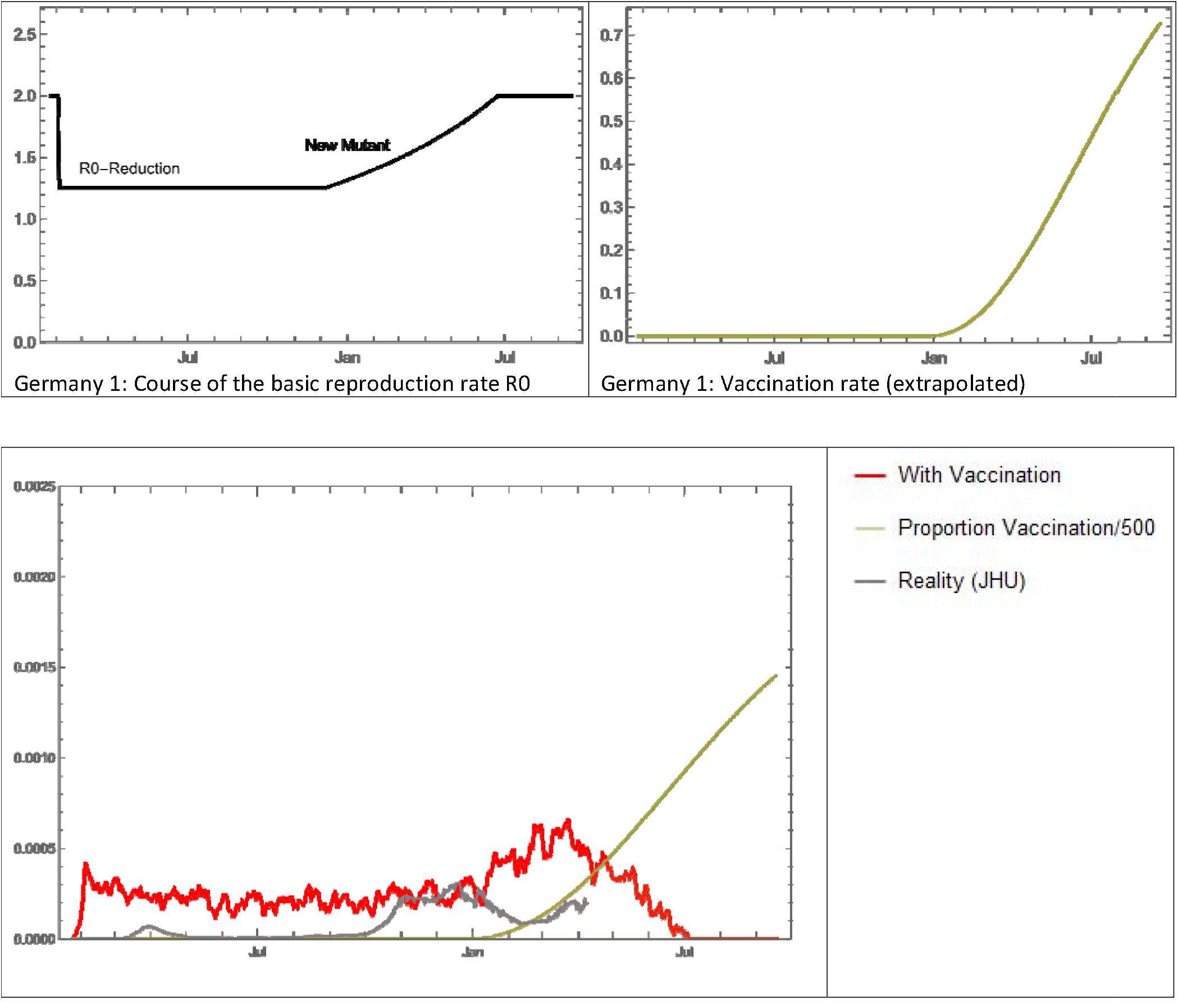
Germany 1: The red curve shows the simulation of the infection with vaccination, the grey curve shows the real course of the infection (Moving Average 7 Days), green: schematic the vaccination rate.

**Fig.3d.**
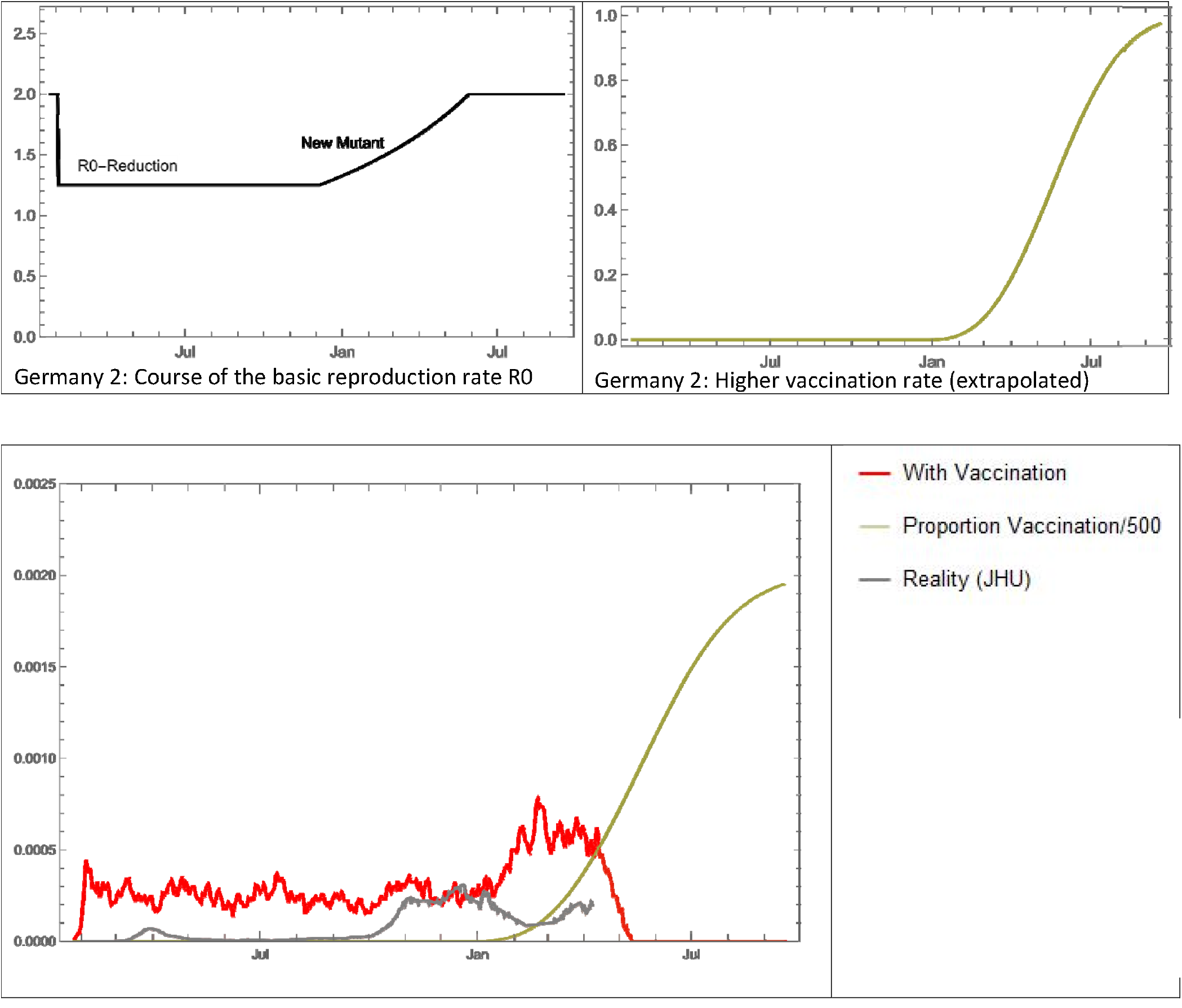
Germany 2 The red curve shows the simulation of the infection with vaccination, the grey curve shows the real course of the infection (Moving Average 7 Days), green: schematic the vaccination rate.

**Fig.3e.**
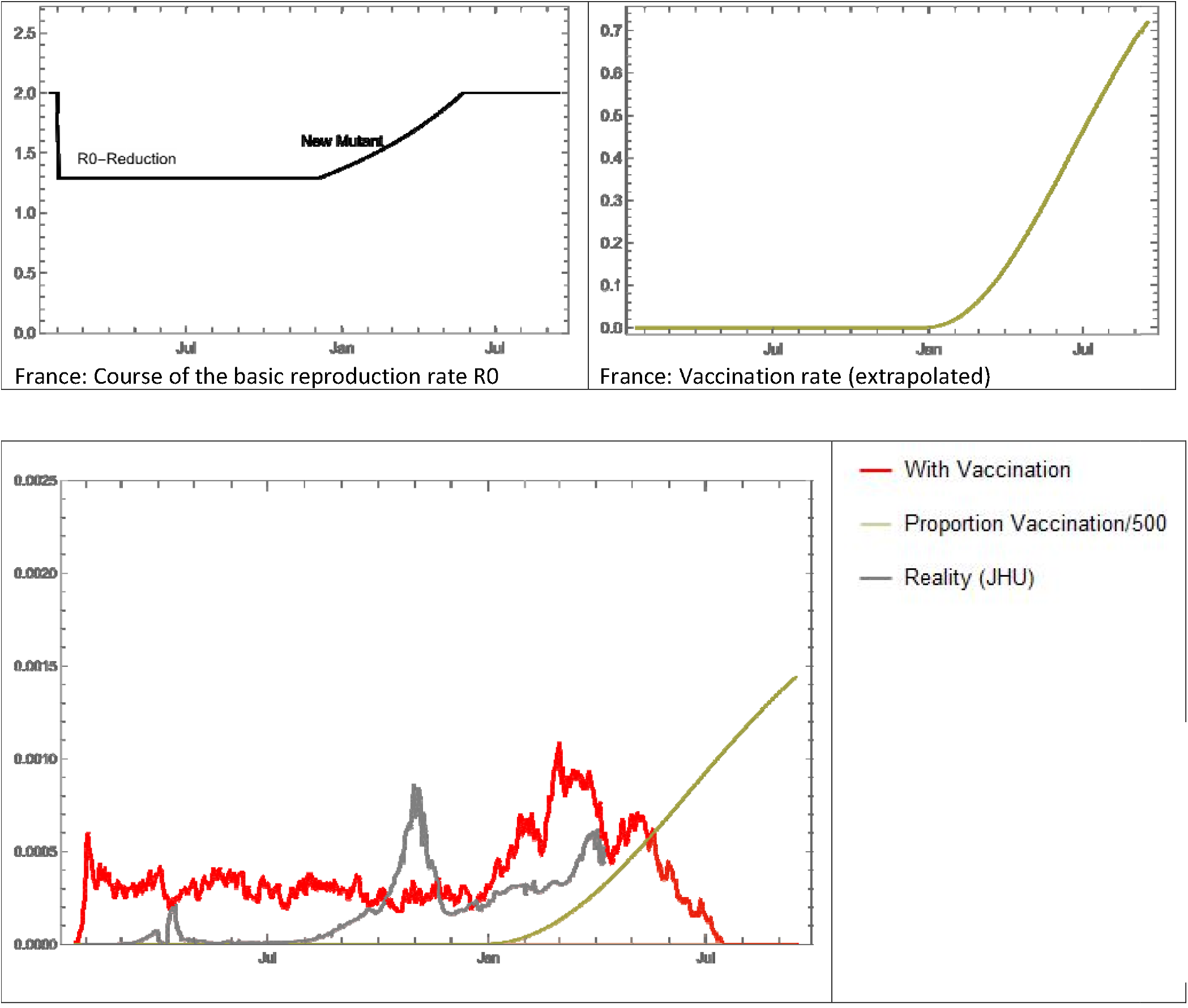
France: The red curve shows the simulation of the infection with vaccination, the grey curve shows the real course of the infection (Moving Average 7 Days), green: schematic the vaccination rate.

**Fig.3f.**
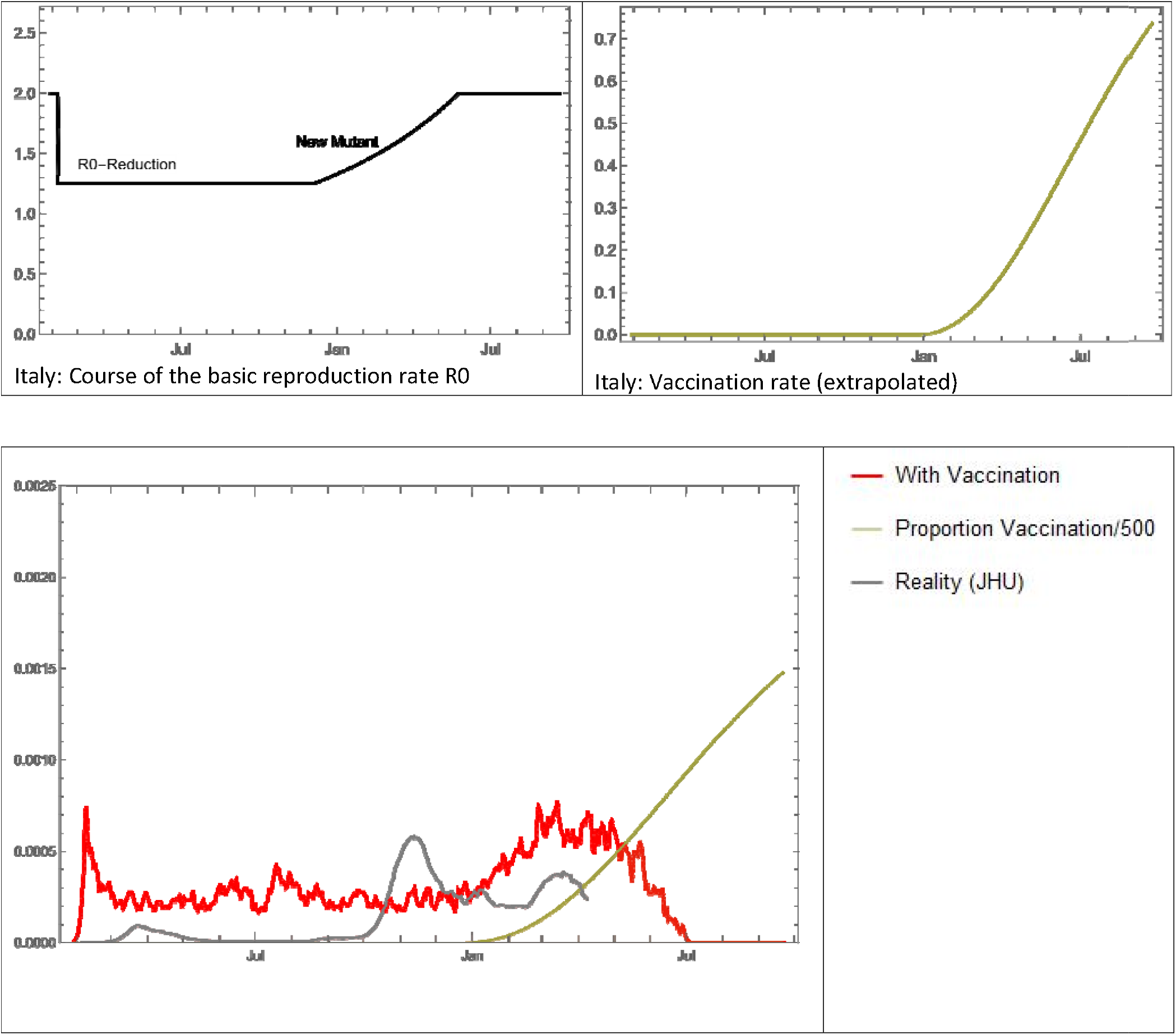
Italy: The red curve shows the simulation of the infection with vaccination, the grey curve shows the real course of the infection (Moving Average 7 Days), green: schematic the vaccination rate.

**Fig.3g.**
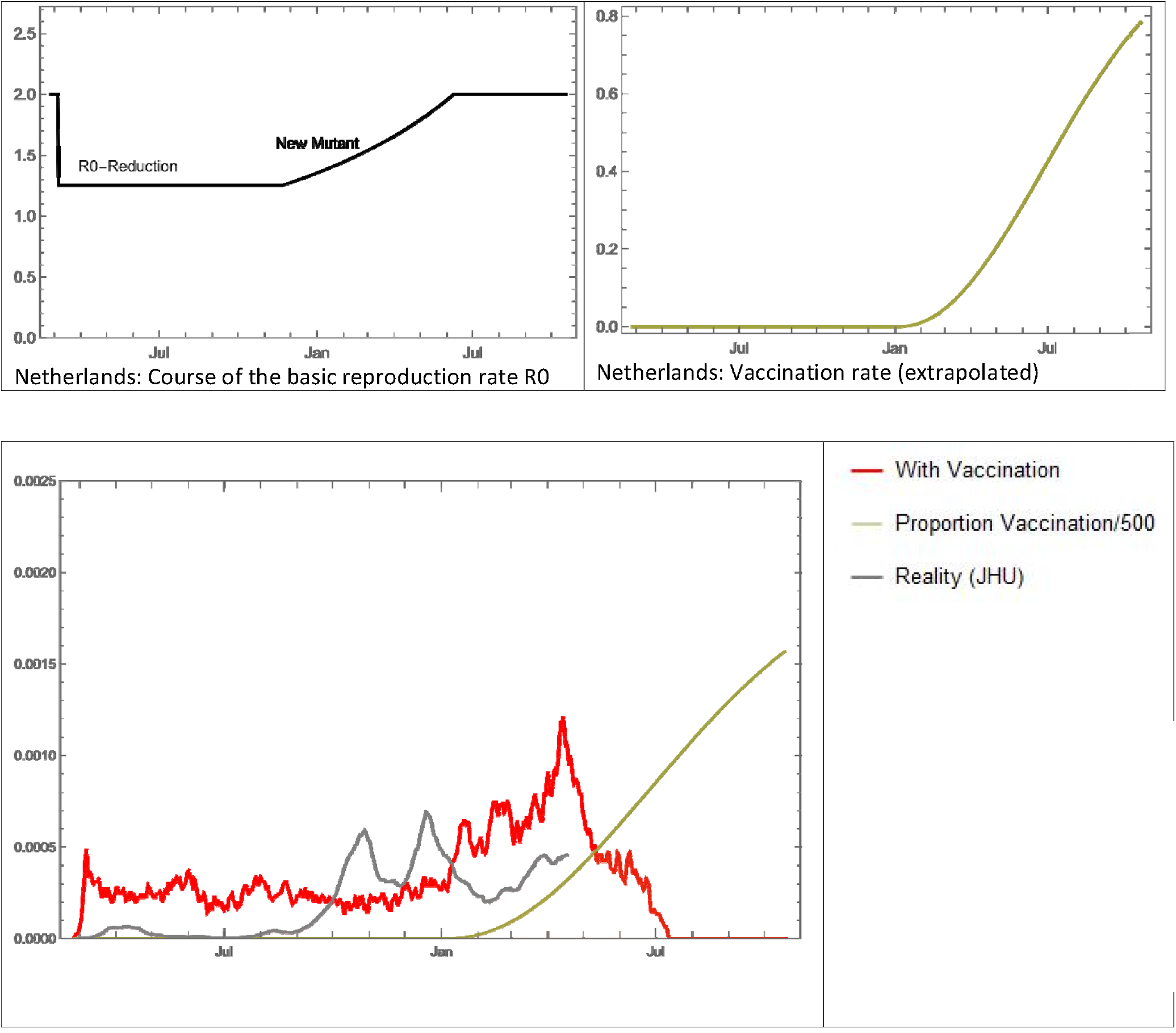
Netherlands: The red curve shows the simulation of the infection with vaccination, the grey curve shows the real course of the infection (Moving Average 7 Days), green: schematic the vaccination rate.

It can be seen that through the corresponding calibration, the courses for the UK and Israel are reproduced surprisingly well and in particular the end of the infection in the simulation corresponds well with the real values. With regard to the total number of infected persons at the end of the infection, there is also a surprisingly good agreement for these two countries: Table 1, last column.

In the simulation for Germany (Fig. 3c) with the real vaccination rate, there are clear deviations from January 2021 onwards, which are probably due to the lockdown at the end of 2020. Under the assumed conditions, the infection comes to a standstill in the simulation in early to mid-July. In another scenario for Germany, a higher vaccination rate was assumed. It was assumed that 45 percent of the population is vaccinated by 15-May-2021. This significantly shortens the end of infection by about 7 weeks in the simulation. Simulations were also calculated for France, Italy and the Netherlands with the results listed in Table 1.

## Discussion

There are now a large number of simulators and simulations of infection trends, both on the basis of differential equation systems and agent-based models. The advantage of agent-based models is that they can be programmed as a stochastic process, corresponding to a real infection event, which is also a stochastic event. In the relatively simple model used here, no emphasis is placed on the exact modelling of an infection process; rather, the infection process is to be considered in terms of the total number of infected persons and, in particular, the end of an infection. This works surprisingly well for the two countries, the UK and Israel. It remains to be seen to what extent the results of the simulations also apply to the other countries studied. However, vaccination rates in particular have a strong impact on the results of the simulations. If, in a few weeks, the vaccination rates are known from the countries where Covid-19 infection is currently still significantly more active than in the UK and Israel, the model can thus be further verified.

## Data Availability

The program and the data are available via the link provided. Attention: stochastic simulation, delivers random values!

https://www.magentacloud.de/share/sdgblo2l5w

## Notes

### Competing Interest Statement

The authors have declared no competing interest.

### Funding Statement

no external funding was received

